# Sex-dependent Clinical Presentation, Body Image, and Endocrine Status in Long-term Remitted Anorexia Nervosa

**DOI:** 10.1101/2023.02.07.23285485

**Authors:** Louisa Schloesser, Leon D. Lotter, Jan Offermann, Katrin Borucki, Ronald Biemann, Jochen Seitz, Kerstin Konrad, Beate Herpertz-Dahlmann

**Affiliations:** Child Neuropsychology Section, Department of Child and Adolescent Psychiatry, Psychosomatics and Psychotherapy, University Hospital RWTH Aachen, Germany; Department of Child and Adolescent Psychiatry, Psychosomatics and Psychotherapy, University Hospital RWTH Aachen, Germany; Institute of Neuroscience and Medicine, Brain & Behaviour (INM-7), Jülich Research Centre, Germany; Institute of Systems Neuroscience, Medical Faculty, Heinrich Heine University Düsseldorf, Germany; Max Planck School of Cognition, Stephanstrasse 1A, 04103 Leipzig, Germany; Institute for Clinical Chemistry and Pathobiochemistry, Otto von Guericke University Magdeburg, Germany; Institute for Laboratory Medicine, Clinical Chemistry and Molecular Diagnostics, University Hospital Leipzig, Germany; JARA-Brain Institute II, Molecular Neuroscience and Neuroimaging, Jülich Research Centre, Germany

**Keywords:** Anorexia nervosa, sex, remission, hormones, body image

## Abstract

**Objective:** Although anorexia nervosa (AN) in males has recently gained attention, knowledge of its psychological and physiological outcomes is still scarce. We explore sex-specific characteristics of long-term remitted AN with respect to residual eating disorder psychopathology, body image, and endocrinology.

**Method:** We recruited 33 patients with AN in remission for at least 18 months (24 women, 9 men) and 36 matched healthy controls (HCs). Eating disorder psychopathology and body image ideals were assessed via clinical interviews, questionnaires, and an interactive 3D body morphing tool. Plasma levels of leptin, free triiodothyronine, cortisol, and sex hormones were quantified. Univariate models controlled for age and weight were used to test for the effects of diagnosis and sex.

**Results:** Both patient groups showed residual eating disorder psychopathology but normal weight and hormone levels relative to HCs. Male remitted patients demonstrated significantly stronger muscularity-focused body image ideals, evident in interviews, self-reports, and behavioural data, than both female patients and HCs.

**Conclusions:** Sex-specific body image characteristics in patients with remitted AN point towards the need to adjust test instruments and diagnostic criteria to male-specific psychopathology. In the future, sufficiently powered studies should evaluate the risk of men with AN developing muscle dysmorphia in the long term.

**Highlights:**

- Gender-specific residual symptoms have been identified in long-term remitted AN patients, particularly regarding body image disturbance.
- The drive for muscularity in male remitted patients suggest a possible risk of transition from male AN to muscle dysmorphic disorder.
- Gender-specific criteria, especially muscle-oriented behaviour, need to be included in the description of diagnoses and symptoms in male patients.

## Introduction and Aims

Anorexia nervosa (AN) is a severe psychiatric disorder with significant morbidity and mortality that predominantly affects female adolescents and young adults (Herpertz-Dahlmann, 2015). Clinical features include severely restricted food intake leading to or accompanied by low body weight, severe fear of weight gain, and body image disturbance (American Psychiatric Association, 2013). With male-female ratios estimated at approximately 1 to 10–20 and a life-time prevalence of 0.16 to 0.3% (van Eeden et al., 2021), occurrence of AN in boys and men was long considered an “atypical” manifestation of the disorder (Murray, Nagata, et al., 2017). Historically and socially shaped stereotypes, as well as differing clinical presentations, may have further hindered significant improvements in AN diagnosis and treatment in boys and men (Zhang, 2014). Indeed, with the DSM-IV, male patients often did not fulfil the criteria for a “full” AN diagnosis and were diagnosed with an “eating disorder not otherwise specified” (ED-NOS), potentially excluding them from both appropriate treatment and contribution to research findings (Raevuori et al., 2014). Given that the numbers of diagnosed male patients have increased since the removal of the EDNOS category and the redefinition of AN diagnostic criteria in the DSM-5 (Vo et al., 2016), and considering increasing awareness and improved knowledge of the clinical presentations of AN in males, sex ratios might adapt in future surveys.

To date, the few clinical trials in acute AN that included male patients and explicitly analysed sex differences showed comparable comorbidity profiles, clinical histories, and behavioural symptoms but more pronounced weight and shape concerns in females with AN (Gorrell et al., 2021). While caloric restriction was the most common strategy to achieve weight loss irrespective of sex, male patients seem to engage more strongly in excessive exercising (Silla et al., 2021). The short-term clinical outcome of AN in males was reported to be better than that in females (Coelho et al., 2021). However, the long-term courses were described to be comparable in both sexes, although previous research emphasized the need for a broader range of assessed outcomes in males (e.g., drive for muscularity) (Quadflieg et al., 2022). From research findings that were primarily obtained in female patients with AN (Tomba et al., 2019), even in a long-term recovered state and in the absence of any fulfilled DSM-5 criteria, significant residual eating disorder (ED) psychopathology can also be expected for males (Quadflieg et al., 2022). However, the very limited data on potential specific psychopathological and biological characteristics of long-term outcomes of AN in males remitted from AN requires further evaluation.

Consistent with the sex-specific clinical presentations in the form of lower weight phobia and shape concerns but greater excessive exercising in male patients, the body image ideals of patients with acute AN might also differ between sexes. While in females with AN body image disturbance is characterized by an overestimation of one’s own body weight and an insatiable desire for thinness (Dalhoff et al., 2019; Provenzano et al., 2019), male patients generally present with a strong drive for muscularity and leanness (i.e., low body fat), while low body weight itself is often considered secondary (Gorrell & Murray, 2019). In female patients, as with ED psychopathology, characteristics in perception of body image disturbance often, but not always, improve with weight gain (Dalhoff et al., 2019; (Eshkevari et al., 2014).

Usually, body image disturbance in AN is assessed using ED-specific interviews and questionnaires, often developed and validated primarily in female populations (e.g., Fairburn et al., 2014). However, muscularity-focused body image ideals, potentially present in males with AN, may be better captured by the *drive for muscularity* concept (McCreary & Sasse, 2000), designed as a measure of male body image ideals. Additionally, computer-based body modelling tools have been increasingly applied in AN showing, for example, greater inaccuracy in perception when modelling one’s current body and significantly distorted body image ideals if asked to model one’s desired body (Mölbert et al., 2018; Ralph-Nearman et al., 2021). While deviant body image ideals are generally well characterized in females with AN, little is known about whether these ideals, and especially their sex-specific variations, occur only in the acute state of the disorder or whether they persist into the long-term course of the disease in both sexes.

Physiologically, eating behaviour is influenced by a variety of hormones that are in turn altered by weight and AN disease status. Indeed, alterations in endocrinological systems can serve as a marker of disease severity and rehabilitation status (Schorr & Miller, 2017). However, data on hormonal status in males with AN, especially in the clinically remitted state, are scarce. During the acute phase of AN, hormonal alterations in free triiodothyronine (fT3), luteinizing hormone (LH), follicle-stimulating hormone (FSH), and leptin, among others, were reported in both men and women (Skolnick et al., 2016; Støving, 2019; Toifl et al., 1988). Compared to healthy controls of similar pubertal status, men with acute AN showed lower total testosterone levels (Schorr & Miller, 2017; Silla et al., 2021). In female patients, most endocrine disturbances improve after weight restoration (Baskaran et al., 2017; Schorr & Miller, 2017). With the exception of a series of three male cases that showed a similar pattern of endocrinological recovery during weight gain to that previously shown in underweight females (Wabitsch et al., 2001), potential hormonal alterations in male patients remitted from AN have not yet been systematically studied.

In this study, we aimed to identify sex-specific residual characteristics of patients with AN in long-term remission using an explorative study design including male and female patients. To quantify residual ED psychopathology and distorted body image, we acquired interview and self-report data and determined aberrant drive for muscularity and body image ideals using a computer-based 3D modelling tool. Hormonal status was assessed as a marker of biological rehabilitation. Based on the limited literature available to date, we expected significant residual ED psychopathology despite normalized weight in AN patients relative to controls, accompanied by potential sex differences concerning body image ideals.

## Methods

### Study sample

The study sample consisted of N = 33 patients remitted from AN, n = 24 women (fAN) and n = 9 men (mAN), and 36 healthy control subjects (fHC, mHC), matched for sex, body mass index (BMI), age, and verbal intelligence (Table 1). “Sex” was defined based on clinical records, and no subject reported a transgender identity. Sexual orientation was assessed via three self-report questions (“To what extent do you consider yourself heterosexual/homosexual/bisexual?”). Patients and male HCs were of central European origin, while 3 female HCs were of Asian origin. All patients (i) had a lifetime history of AN according to DSM-5 criteria (American Psychiatric Association, 2013) and (ii) had received inpatient, day-care or outpatient treatment for AN. HC subjects were included if they (i) had a BMI higher than 18 kg/m^2^ and (ii) had no current or previous psychiatric or neurological disorders as assessed by a screening interview. N = 3 subjects (n = 2 fAN, n = 1 mAN) were excluded from the initial cohort of 36 patients because they fulfilled the diagnostic criteria for bulimia nervosa (BN).

**Table 1:** Demographic and clinical sample data. Variables were compared between groups as appropriate, using two-way analyses of variances with bootstrapping followed by evaluation of simple main effects, Mann-Whitney U tests, or Chi-squared tests. Missing values: a: n = 1 fHC, 1 mHC. b: n = 1 fAN, 1 mAN. c: n = 1 fHC. d: n = 1 fAN, 1 mAN, 3 fHC, 1 mHC. e: n = 3 fAN, 1 mAN, 5 fHC, 4 mHC. f: n = 2 fAN, 1 mAN, 1 fHC, 2 mHC. g: n = 1 fAN, 1 mAN, 1 fHC, 2 mHC. SD = standard deviation, y = years, IQ = intelligence quotient, BMI (-SDS) = body mass index (-standard deviation score), EDE = Eating Disorder Examination, EDI-2 = Eating Disorder Inventory, BDI-2 = Beck Depression Inventory 2, BAI = Beck Anxiety Inventory, MINI = Mini International Neuropsychiatric Interview, M/m = male, F/f = female, ns. = not significant.

|  | Remitted Anorexia nervosa (AN) |  | Healthy controls (HC) |  | Comparison |
| --- | --- | --- | --- | --- | --- |
|  | Female (N = 24) | Male (N = 9) | Female (N = 26) | Male (N = 10) |  |
|  | Mean [SD] / N, Range | Mean [SD] / N, Range | Mean [SD] / N, Range | Mean [SD] / N, Range |  |
| *Age (y)* | 22.0 [3.2], 17.7 – 31.4 | 24.1 [4.4], 18.7 – 32.5 | 22.5 [3.5], 16.6 – 31.3 | 25.6 [3.7], 20.3 – 31.0 | M>F |
| *Verbal IQ<sup>a</sup>* | 106.3 [10.4], 85 – 124 | 104.8 [12.9], 92 – 136 | 110.6 [11.8], 94 – 130 | 107.8 [7.2], 95 – 118 | ns. |
| *BMI (kg/m<sup>2</sup>)* | 21.3 [2.7], 18.1 – 26.6 | 22.9[3.5], 18.3 – 27.4 | 21.6 [2.1], 18.2 – 25.5 | 23.1 [1.9], 19.4 – 25.5 | M>F |
| *BMI-SDS* | -.4 [.9], -1.7 – .9 | -.4 [1.1], -2.2 – .8 | -.3 [.7], -1.6 – .9 | -.4 [.6], -1.4 – .5 | ns. |
| *Age at first admission (y)<sup>b</sup>* | 15.0 [1.5], 12.0 – 18.0 | 15.1 [1.9], 12.4 – 18.1 | - | - | ns. |
| *Treatment duration (y)<sup>b</sup>* | 1.7 [1.1], .1 – 4.4 | 2.3 [2.2], .1 – 6.7 | - | - | ns. |
| *Post-treatment duration (y)<sup>b</sup>* | 5.2 [2.7], 1.9 – 13.9 | 7.4 [4.5], 2.7 – 16.3 | - | - | ns. |
| *EDE – mean total score* | 1.0 [.9], .1 – 3.7 | .5 [.3], .1 – 1.1 | - | - | ns. |
| *EDI-2 – total score<sup>c</sup>* | 260.1 [70.9], 175 – 399 | 213.6 [56.2], 150 – 321 | 193.0 [35.8], 151 – 263 | 173.3 [21.6], 143 – 222 | AN>HC, W>M, fAN>fHC, mAN>mHC |
| *BDI-2<sup>d</sup>* | 11.3 [12.0], 0 – 37 | 7.8 [8.5], 0 – 26 | 5.2 [4.7], 0 – 14 | 3.3 [3.7], 0 – 10 | AN>HC, fAN>fHC |
| *BAI<sup>e</sup>* | 6.7 [7.1], 0 – 29 | 4.8 [4.3], 2 – 13 | 4.4 [5.8], 0 – 24 | 1.3 [1.0], 0 – 3 | ns. |
| *SF-36 – total score<sup>f</sup>* | 84.6 [14.2], 42.5– 114 | 84.7 [8.4], 68.6 – 94.6 | 83.9 [7.9], 62.5 – 95.4 | 91.4 [4.0], 84.9 – 96.4 | ns. |
| *SF-36 – mental health<sup>g</sup>* | 79.6 [17.4], 35 – 100 | 71.9 [22.0], 35 – 95 | 80.2 [13.4], 50 – 100 | 85.9 [10.8], 72 – 100 | ns. |
| *Heterosexuality* | 6.7 [1.0], 2 – 7 | 7 [0], 7 | 6.1 [1.8], 1 – 7 | 6.9 [0.3], 6 – 7 | ns. |
| *Homosexuality* | 1 [0], 1 | 1 [0], 1 | 1.0 [0.2], 1 – 2 | 1 [0], 1 | Ns. |
| *Bisexuality* | 1.2 [0.4], 1 – 2 | 1 [0], 1 | 2.1 [1.6], 1 – 6 | 1.1 [0.3], 1 – 2 |  |
| *Hormonal contraception* | 10 | - | 19 | - | fHC>fAN |
| *MINI – $\geq 1$ comorbidity* | 8 | 1 | - | - | ns. |

The study was approved by the local ethics committee (EK 213/15) and was conducted in accordance with the Declaration of Helsinki, and all participants and their legal guardians (if underage) gave written informed consent. Former patients who received inpatient treatment with a diagnosis of AN were recruited from the University Hospital RWTH Aachen, Germany, and surrounding clinics specializing in the treatment of eating disorders. HC subjects were recruited via postings from the surroundings of the University Hospital RWTH Aachen without being informed about the goals of the study. All participants received financial compensation in accordance with the local standards for clinical studies.

The data and results reported here were published in part in LS’s medical thesis (Schlösser et al. (2022); neuroimaging data collected from an overlapping sample (n = 21 female patients and N = 22 female HCs) were published in another report (Lotter et al., 2021).

### Clinical characteristics and psychopathology

In patients, data on the current state and long-term clinical course of EDs were collected in structured interviews and from medical records if the latter were available (n = 31 patients). *Treatment duration* was defined as the time between first admission and last discharge from inpatient and/or outpatient treatment with a diagnosis of AN; *posttreatment duration* was defined as the time between last discharge and the study date. Body weight was measured with a standardized scale in the morning prior to food intake and adjusted for height, age, and sex by calculation of the BMI–standard deviation score (BMI-SDS) (Hemmelmann et al., 2010; Neuhauser et al., 2013). ED diagnostic criteria and severity were assessed using the Eating Disorder Examination (EDE) and the Eating Disorder Inventory 2 (EDI) (Fairburn et al., 2014; Garner, 1991). EDE criteria for diagnosis of AN and BN according to DSM-5 were selected to assess weight phobia (AN-B: *Fear of gaining weight*), body image disturbance (AN-C: *Disturbance of body weight/shape experience*), binge eating (BN-A: *Recurrent episodes of binge eating*), and “purging” behaviour (BN-B: *Recurrent inappropriate compensatory behaviours to prevent weight gain*). Comorbid psychiatric disorders were screened using the Mini International Neuropsychiatric Interview (MINI) (Ackenheil et al., 1999). The Beck Anxiety Inventory (BAI) (Margraf & Ehlers, 2007) and the revised Beck Depression Inventory (BDI) (Hautzinger et al., 2006) were used to assess anxiety and depressive symptoms. Health-related quality of life was assessed with the 36-Item Short Form Health Survey (SF-36) (Lins & Carvalho, 2016). The Multiple-choice Vocabulary Test (Lehrl, 2005) was utilized to approximate the verbal intelligence quotient. Interviews were conducted by a specifically trained and supervised clinical researcher (JO).

To be considered “remitted”, patients had to meet the following criteria: (i) no fulfilment of full ED criteria according to the Eating Disorder Examination (EDE) (Fairburn et al., 2014), (ii) a BMI of at least 18 kg/m^2^ or, if underage, at least the 10^th^ percentile, (iii) no current treatment for AN, (iv) no report of ED-related behaviour for at least 1.5 years, and (vi) a stable weight in the time period since the last inpatient or outpatient discharge. Considering the post-treatment time of 5.5 years on average, we adopted the term *long-term remitted*.

### Body image and drive for muscularity

Characteristics of body image disorder were assessed using 3D modelling software (“Body Morphing Tool” (BMT), DAZ Studio 4 Pro, Daz3D.com) (Crossley et al., 2012). Participants were seated in front of a 15-inch notebook and asked to model their own body (i) as they perceived it at the time of testing and (ii) as they imagined their “desired” body. The avatars were sex-specific and could be modified by slide controls in five categories selected from the software’s presets: “Body Size”, “Weight” (ranging from -100 to +100%), *“*Bodybuilder”, *“*Fitness”, and “Stocky” in males or “Voluptuous” in females (ranging from 0 to 100%). The BMT allowed free rotation and zooming into the body (Figure S1). Values corresponding to the slide control positions of the categories “Body builder”, “Fitness”, and “Weight” were used for group comparisons (hereafter referred to as *muscularity, definition*, and *body fat*). From the five groupwise mean BMT indices, we created visualizations of the “average” perceived and desired bodies for each group. To assess muscularity-related attitudes and behaviour, we applied the Drive for Muscularity Scale (DMS), consisting of 15 questions (Waldorf et al., 2014).

### Hormonal status

Blood was drawn in the morning after an overnight fast, centrifuged, aliquoted, and stored at -80 °C. Concentrations of leptin, free triiodothyronine, cortisol, LH, FSH, progesterone, estradiol, and testosterone were determined by enzyme-linked immunosorbent assays (laboratory of the University Hospital Magdeburg, Germany). Subjects using hormonal contraception were excluded from the analyses of FSH, LH, progesterone, oestrogen, and testosterone because of its influence on sex hormone levels.

### Statistical analyses and visualizations

Statistical tests were carried out using SPSS (IBM Corp., 2020, version 26). Due to multiple nonnormally distributed variables as evaluated using Shapiro-Wilk tests and the small sample sizes of the male groups, nonparametric statistics and bootstrapping procedures were used whenever possible. Because of the small group sizes, EDE diagnostic criteria were compared between groups using two-sided Fisher’s exact tests. Demographic, clinical, and self-report data (EDI subscales, EDE, BAI, BDI, SF-36, DMS), hormonal plasma concentrations, and parameters derived from the BMT were compared using two-way analyses of covariance (ANCOVAs) with diagnosis and sex as main factors. Age and BMI-SDS were included as covariates in all models. Main and diagnosis × sex interaction effects were evaluated and followed up on by evaluation of the simple main effects of diagnosis within sex categories (Šidák correction). Bootstrapping procedures as implemented in SPSS (N = 1000 samples; random number generator: Mersenne Twister, seed: 2000000) were used in ANCOVA models to account for violations of normality assumptions (Chernick, 2008).

In sensitivity analyses, we repeated all ANCOVAs while (i) excluding all subjects with a BMI below the 10^th^ age- and sex-adjusted percentile, (ii) including three additional subjects with current BN, (iii) applying binary logistic regression to control significant group comparisons of EDE criteria for the influence of BMI-SDS and age, and (iv) using Mann-Whitney U (MWU) tests to assess significant ANCOVA results for the potential impact of outliers. Correlational analyses were conducted separately within the respective subgroups using Spearman correlations. Due to the exploratory nature of this study and the assessment of multiple inter-correlated variables, the significance level was set at two-tailed α = .05 and Bonferroni-corrected per set of analyses (in the case of ANCOVAs, applied to assess the significance of the complete model): EDE criteria (4 tests), EDI subscales (11 tests), hormone levels (8 tests), and drive for muscularity/BMT (7 tests).

Visualizations were created using ggplot2, version 3.3.5, (Wickham, 2016) in R, version 3.6.3, (R Core Team, 2021).

## Results

### Sample characteristics and general results

Age differed between males and females but not between former patients and HCs. No group differences were found for verbal intelligence, BMI, or BMI-SDS. Sexual orientation was largely heterosexual and did not differ between groups (n = 2 fHC: bisexual orientation, n = 1 fHC: no sexual preference). Female and male patients were comparable regarding age at admission, treatment and posttreatment durations, EDE total scores, and current number of comorbidities. Patients presented with higher overall ED, depression, and anxiety scores than HCs, but the groups did not differ regarding quality of life (Table 1, Table S1).

### Residual ED psychopathology

When applying Bonferroni correction (corrected α =.0125), EDE criteria frequency did not differ between male and female patients remitted from AN. However, numerically, more male than female patients fulfilled the EDE criterion BN-B: *Recurrent inappropriate compensatory behaviours to prevent weight gain*, although the result was not significant (χ^2^(1) = 7.64, p =.013, uncorrected), while the opposite was the case for AN-C: *Disturbance of body weight/shape experience* (χ^2^(1) = 4.91, p =.044, uncorrected), being observed more frequently in females. Further analyses revealed that compensatory behaviours consisted of pathologically driven exercising in all mAN subjects. The other criteria did not differ between the sex groups (AN-B: χ^2^(1) =.39, p = 1.000; BN-A: χ^2^(1) =.76, p =.642; Figure 1A). Bootstrapped binary logistic regression analyses of criteria BN-B and AN-C indicated robustness against influences of age and BMI-SDS (BN-B: χ^2^(3) = 7.89, p =.048, Sex: B = -2.39, SE = 7.27, p =.010; AN-C: χ^2^(3) = 5.11, p =.164, Sex: B = -1.94, SE = 6.92, p =.013).

**Figure 1:**
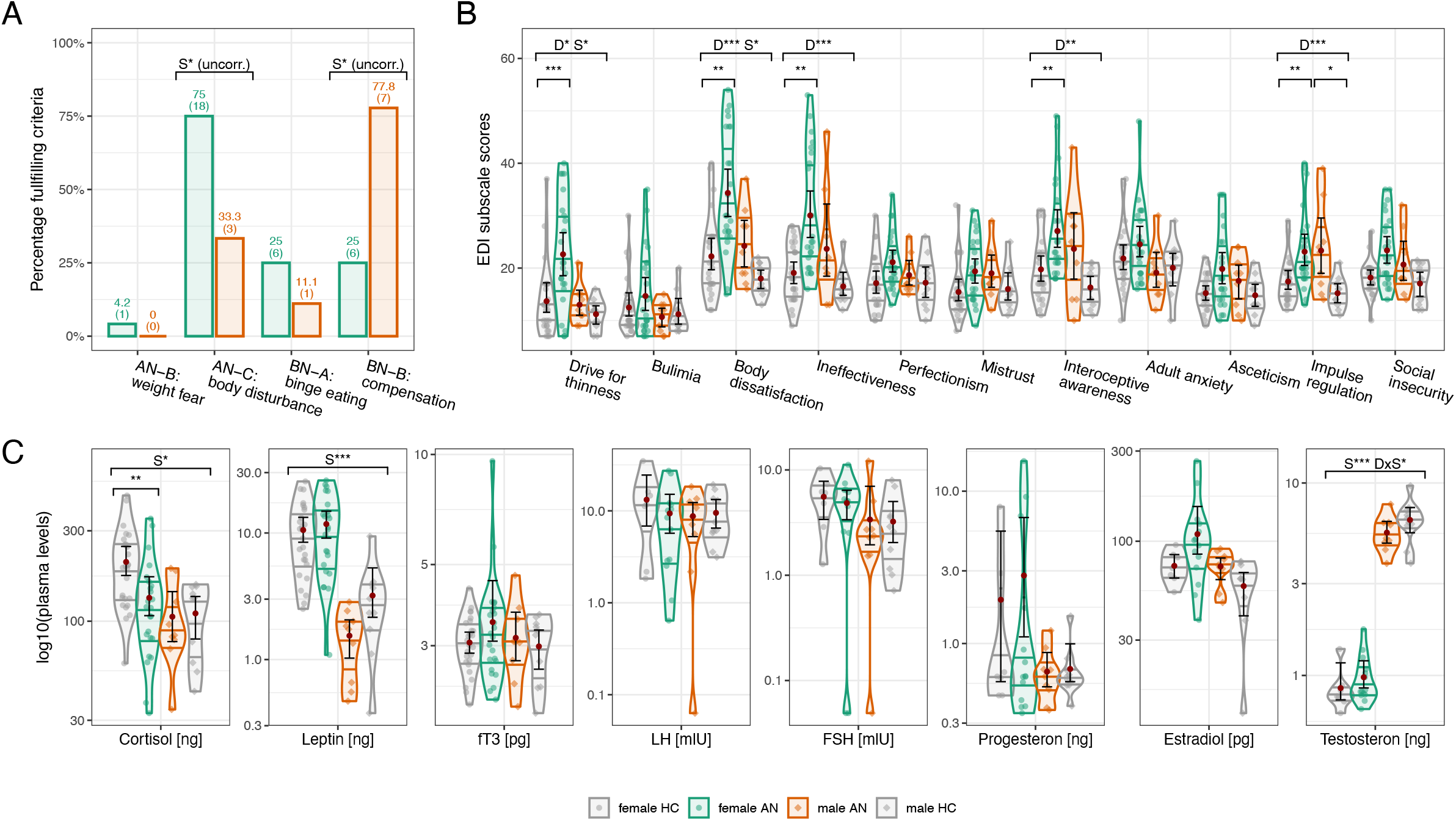
Residual eating disorder psychopathology and hormonal status. **A**: Percentages of female and male patients fulfilling the preselected diagnostic criteria for AN (AN-B: *Fear of gaining weight*, AN-C: *Disturbance of body weight/shape experience*) and BN (BN-A: *Recurrent episodes of binge eating*, BN-B: *Recurrent inappropriate compensatory behaviours to prevent weight gain*). Numbers in bars: “percent (n)”. **B** and **C**: Group-wise results of EDI subscales and hormone assessments, respectively. Hormone levels are given per milliliter and log10-scaled. Individual values are shown as scatter points and squares, violins depict distributions of values within each group (not scaled to group size but to same width per group), red circles and error bars show group-wise mean values and corresponding bias-corrected and accelerated (BCa) bootstrap confidence intervals. Brackets show significant group comparison results, * p < .05, ** p < .01, *** p < .001, D = effect of diagnosis, S = effect of sex, DxS = interaction effect. AN = Anorexia nervosa, BN = Bulimia nervosa, HC = Healthy control, EDI = Eating Disorder Inventory 2, fT3 = free triiodothyronine, LH = luteinizing hormone, FSH = follicle-stimulating hormone. See Tables S2 and S3 for detailed results.

Bootstrapped two-way ANCOVA models of the EDI subscales were significant for the scales *drive for thinness, body dissatisfaction, ineffectiveness, interoceptive awareness*, and *impulse regulation*. In all cases, we found significant main effects of diagnosis, with AN patients showing higher scores than HCs with large effect sizes. A significant main effect of sex revealed a higher *drive for thinness* and *body dissatisfaction* in females than in males. The analyses of simple main effects further indicated that the effects of diagnosis were strongly driven by the group differences between fAN patients and fHCs; however, no interaction effects emerged. When applying Bonferroni correction, no model was significant, but some main effects emerged for the remaining subscales (Table S3, Figure 1B). Recalculating the significant models while (i) excluding participants with a BMI below the 10^th^ percentile, (ii) including subjects with current BN, and (iii) using MWU tests confirmed the stability of the group effects (Tables S4–S6).

### Body image and drive for muscularity

ANCOVAs assessing the BMT-derived variables *desired muscularity* and *desired* and *perceived muscle definition* were significant on the model level and showed significant sex, diagnosis × sex interaction, and, except for *desired muscle definition*, diagnosis effects (Bonferroni corrected, large effect sizes). For *perceived muscularity*, we observed effects of the diagnosis main factor and the interaction (uncorrected). *Body fat* indices did not differ between groups. Regarding self-reported drive for muscularity (DMS), significant effects of diagnosis and sex emerged. In all cases, AN patients exhibited increased scores relative to HCs, and males showed higher scores than females. Simple main effects indicated a general pattern of higher drive for muscularity in mAN patients than in mHCs that was not present in female subgroups (Table S3, Figure 2A). Group difference patterns largely remained identical when recalculating analyses while applying the stricter BMI criterion, including BN subjects, or using MWU tests (Tables S4–S6). Figure 2B shows groupwise “average” avatars for the *perceived* and *desired* conditions.

**Figure 2:**
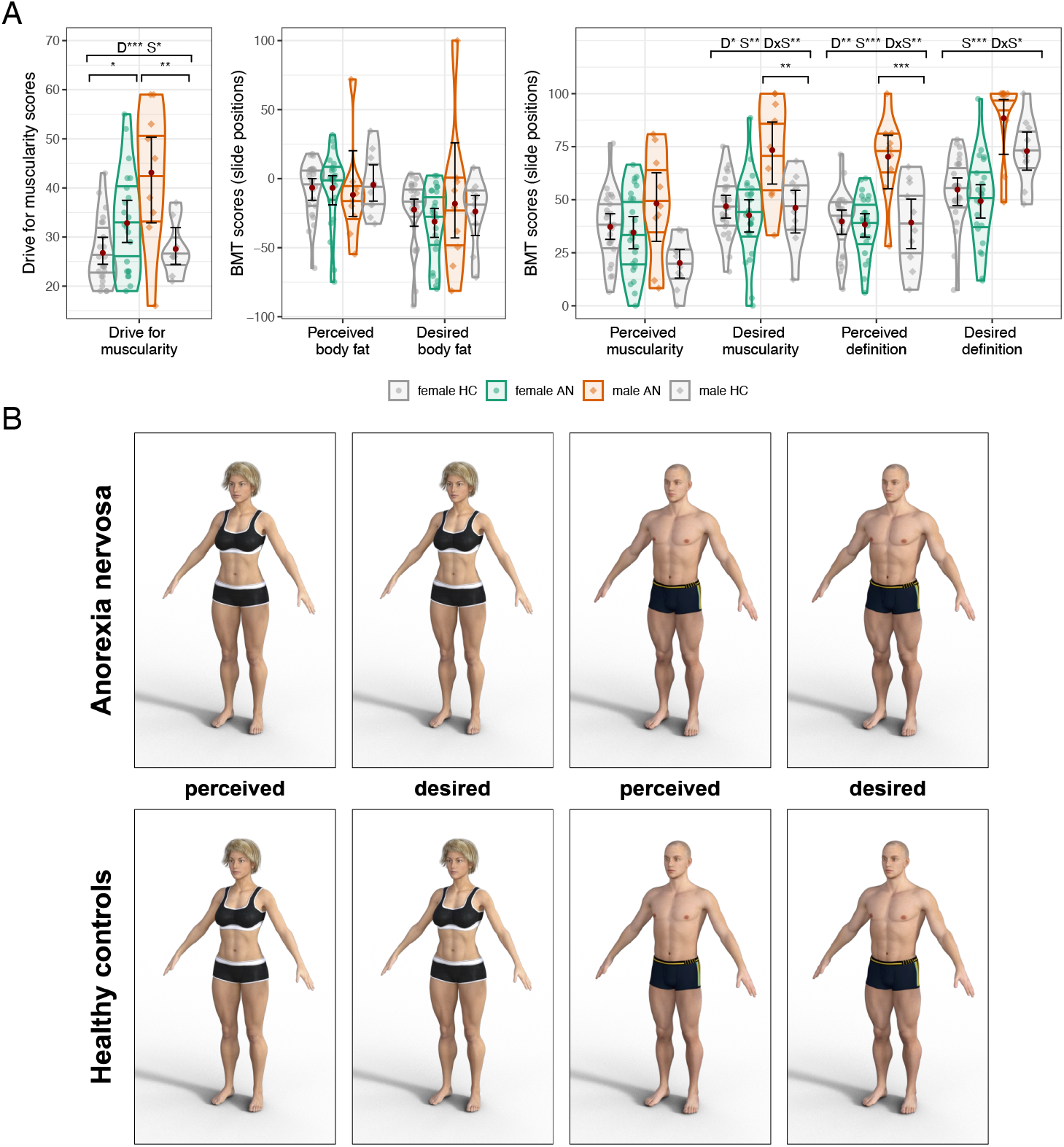
Body image characteristics. **A**: Group-wise results of Drive for Muscularity Scale and Body Morphing Tool (BMT) parameters. See Figure 1 for descriptions. **B**: Group-wise averaged perceived and desired avatars, derived by manually configuring avatars from mean values of five BMT scales and rendering within *DAZ Studio 4 Pro*.

### Hormonal status

ANCOVA models were significant only for leptin, cortisol, and testosterone (Bonferroni corrected; Table S3, Figure 1C). In all cases, plasma levels were significantly affected by sex, with females showing higher levels of leptin and cortisol and males demonstrating higher testosterone levels. For testosterone, we additionally observed a significant interaction between diagnosis and sex with a medium effect size. This effect emerged from reduced testosterone levels in the male AN group but increased testosterone levels in the female AN group compared to sex-specific HC groups. Note, however, that the simple main effects were not significant and that the effect size for the interaction effect was medium. Sensitivity analyses showed no relevant influence of a more conservative BMI cut-off or the additional inclusion of BN subjects on all group effects and a general stability of results against influence of outliers (Tables S4– S6).

### Correlation analyses

Leptin correlated strongly with BMI-SDS in female subgroups but not in males; testosterone and BMI did not show significant associations in any subgroup (Figure S2). Evaluating associations between BMT indices, DMS, *drive for thinness*, and body composition, we found (i) patterns of positive associations between *perceived body fat* and BMI-SDS as well as between leptin and *drive for thinness*, especially in female subgroups, (ii) negative associations between *desired body fat* and DMS scores across groups, and (iii) a strong correlation between perceived muscularity and BMI-SDS only in males with AN (Table S7, Figure S3).

## Discussion

In this exploratory study, we investigated ED psychopathology, body image ideals, and hormonal status in female and male patients with long-term remission from AN. We found significantly elevated ED psychopathology focused on body dissatisfaction in females as opposed to excessive exercising in males. In line with this finding, male patients remitted from AN pursued significantly muscularity-centred body image ideals compared to females and male controls.

Despite the persistent ED symptomatology, we observed almost full endocrinological rehabilitation in remitted AN which was most likely achieved by regular and sufficient energy intake (Støving, 2019). The observed sex differences in leptin, testosterone, and cortisol levels can be considered physiological (Asarian & Geary, 2013; Christen et al., 2018; Goel et al., 2014). Elevated ED psychopathology in (female) patients in various states of recovery from AN can be considered a well-replicated finding (Bernardoni et al., 2020; Boehm et al., 2016; Cascino et al., 2020), and thus, our results were to be expected. ED psychopathology differing by sex, with females showing stronger concerns about body weight and shape and males exhibiting especially pronounced driven exercising, was previously reported independently in females and males with acute (Gravina et al., 2018; Mairs & Nicholls, 2016) and short-term remitted AN (Heimann et al., 2018; Silla et al., 2021). We extend these results by showing similar patterns in long-term remission of AN in the form of significantly elevated body dissatisfaction (EDI-2) and a higher frequency of body image disturbance in female remitted AN patients but a significantly higher frequency of compensatory behaviours in male remitted AN patients (EDE).

In line with the above findings, using the BMT, we demonstrated body image ideals centred on muscularity and fitness, accompanied by increased muscularity-oriented attitudes and behaviours (DMS), in male patients remitted from AN in a more naturalistic manner, as was previously reported in acute AN (Murray et al., 2016). In contrast, female patients did not differ from controls on muscularity and body fat ratings, irrespective of the *perceived* or *desired* conditions. On the one hand, this could be interpreted as evidence for a normalization of the distorted body image in females remitted from AN. On the other hand, especially considering the persistently elevated ED psychopathology and drive for muscularity observed in the same patients and the interaction effect of diagnosis and sex on BMT scores, a potential repulsive effect of higher muscularity and subsequently higher body weight specifically in female patients might be discussed. Given the preliminary nature of this study, these findings require replication in a larger cohort based on tools validated in both female and male populations.

Considering that all male patients in this study initially had low body weight and were diagnosed with AN, the observed muscularity-focused body image ideals suggest a possible risk of transition to muscle dysmorphic disorder in the long-term clinical course of males with AN. Muscle dysmorphic disorder, also referred to as “reverse anorexia”, is characterized by the desire for a larger and more muscular body accompanied by the belief that one is small and thin, even though individuals are often tall and muscular (Murray et al., 2010). Indeed, recently, a reconsideration of muscle dysmorphic disorder from the perspective of ED pathology (Gorrell & Murray, 2019) and the corresponding adaptation of treatment strategies (Cunningham et al., 2017) have been proposed. Transitions from AN to muscle dysmorphic disorder in male patients have been previously described (Murray, Griffiths, et al., 2017). However, whether these muscle dysmorphic body image distortions actually have negative impacts on patients’ quality of life remains unclear and could not be verified by the present investigation. However, the association between AN and muscle dysmorphic disorder in males calls for further investigation.

### Limitations

First, this exploratory study is limited by the small sample size, especially of the male subgroup, requiring replication in well-powered studies. Second, it is questionable whether measurements and tests developed primarily for female patients with AN adequately assess AN diagnosis and symptoms in males. The EDE appears to have clinical utility in men (Schaefer et al., 2019), although they tend to score lower than females (Darcy et al., 2012). On the other hand, the EDI-2 did not detect sex differences in the present study, which is in line with its previously reported limited reliability in men (Spillane et al., 2004). Third, the usually applied weight criterion of a threshold BMI of 18.5 kg/m^2^ corresponds to the 10^th^ BMI percentile only in females, but not in males, which generally complicates comparison of sexes in AN. Here, we further included participants based on a BMI cut-off of at least 18 kg/m^2^, whereas the DSM-5 defines “significantly low body-weight” with a BMI threshold of 18.5 kg/m^2^. However, our sensitivity analyses using a more conservative BMI cut-off largely confirmed our findings. Fourth, our approach to measuring body image characteristics is limited, as it only assessed self-reported muscularity-oriented cognition and behaviour as well as body image ideals but lacked assessment of actual fat or muscle mass to validate the subjective measures. In addition, the BMT template avatars were white and not individualized, which could limit the validity of the approach, as participants may not identify with the avatars.

## Conclusion

In line with previous studies (Bardone-Cone et al., 2019), we found evidence for sex-specific characteristics of body image disturbance and drive for muscularity in long-term remitted AN indicating a potential association between male AN and muscle dysmorphic disorder. Thus, we recommend addressing sex-specific ED symptomatology, in particular muscularity-oriented body image ideals and behaviours when assessing diagnosis and symptoms in males with AN. Incorporation of sex-specific symptoms, as well as body weight (BMI) thresholds, in AN criteria for diagnosis, remission, and recovery will have to be discussed in the future. This will require further development of appropriate assessment instruments, especially for male patients with AN (Arkenau et al., 2020).

## Data Availability

The data presented in the study are available upon reasonable request from the author.

## Acknowledgements

We thank the staff of the DRK Fachklinik Bad-Neuenahr, the University Medical Center Utrecht, and the University Hospital Vienna for their support in providing patient contacts and clinical data. LDL was supported by the Federal Ministry of Education and Research (BMBF) and the Max Planck Society (MPG), Germany.

## Author Contributions

LS and LDL performed data analyses and wrote the manuscript. KK, JS and BHD designed the overall study. LS, LDL and JO conducted the clinical study and gathered the data under supervision of JS, KK and BHD. KB and RB monitored the hormone assessments. All authors reviewed and commented on the manuscript.

