## Supplements for "Sex-dependent Clinical Presentation, Body Image, and Endocrine Status in Long-term Remitted Anorexia Nervosa"

### Supplementary Tables

| Comorbidity in MINI | Female [% (N)] | Male [% (N)] |
|---|---|---|
| Major depressive disorder | 16.7 (4) | 0 |
| Suicidality | 8.3 (2) | 11.1 (1) |
| Panic disorder | 12.5 (3) | 0 |
| Agoraphobia | 4.2 (1) | 0 |
| Social phobia | 4.2 (1) | 0 |
| Generalized anxiety disorder | 16.7 (4) | 0 |
| Alcohol abuse | 0 | 11.1 (1) |
| At least one comorbidity | 33.3 (8) | 11.1 (1) |
| No comorbidity | 66.7 (16) | 88.9 (8) |

**Table S1: Current comorbid psychiatric disorders in patients assessed using the Mini International Neuropsychiatric Interview (MINI)**

Percentages of full samples.

| | Recovered anorexia nervosa (AN) | | | | | | | | Healthy Controls (HC) | | | | | | | |
|---|---|---|---|---|---|---|---|---|---|---|---|---|---|---|---|---|
| | Female | | | | Male | | | | Female | | | | Male | | | |
| | *mean* | *SD* | *min* | *max* | *mean* | *SD* | *min* | *max* | *mean* | *SD* | *min* | *max* | *mean* | *SD* | *min* | *max* |
| **Residual eating disorder psychopathology (Eating Disorder Inventory 2)** | | | | | | | | | | | | | | | | |
| *Drive for thinness* [a] | 22.6 | 10.4 | 7.0 | 40.0 | 13.0 | 3.8 | 9.0 | 21.0 | 13.7 | 7.0 | 7.0 | 37.0 | 11.2 | 2.9 | 7.0 | 16.0 |
| *Bulimia* [a] | 14.7 | 8.1 | 7.0 | 35.0 | 10.7 | 3.0 | 7.0 | 15.0 | 12.5 | 5.5 | 7.0 | 30.0 | 11.2 | 4.0 | 8.0 | 20.0 |
| *Body dissatisfaction* [a] | 34.3 | 11.4 | 15.0 | 54.0 | 24.2 | 7.2 | 16.0 | 37.0 | 22.2 | 7.7 | 12.0 | 40.0 | 18.0 | 3.0 | 13.0 | 22.0 |
| *Ineffectiveness* | 30.0 | 11.2 | 16.0 | 53.0 | 23.7 | 10.7 | 13.0 | 46.0 | 19.1 | 5.6 | 9.0 | 28.0 | 16.5 | 3.6 | 12.0 | 25.0 |
| *Perfectionism* [b] | 21.1 | 5.3 | 13.0 | 34.0 | 18.7 | 3.7 | 15.0 | 26.0 | 17.1 | 5.3 | 10.0 | 30.0 | 17.2 | 5.0 | 10.0 | 26.0 |
| *Interpersonal distrust* [a] | 19.4 | 6.0 | 9.0 | 31.0 | 19.0 | 5.0 | 12.0 | 29.0 | 15.4 | 5.2 | 8.0 | 32.0 | 16.0 | 4.2 | 11.0 | 25.0 |
| *Interoceptive awareness* [b] | 27.1 | 9.0 | 18.0 | 49.0 | 23.7 | 10.2 | 10.0 | 43.0 | 19.8 | 6.1 | 11.0 | 31.0 | 16.3 | 3.8 | 11.0 | 21.0 |
| *Maturity fears* [a] | 24.5 | 7.2 | 16.0 | 48.0 | 19.1 | 5.3 | 12.0 | 30.0 | 21.8 | 6.1 | 10.0 | 37.0 | 20.1 | 5.2 | 10.0 | 29.0 |
| *Asceticism* [c] | 19.8 | 7.6 | 10.0 | 34.0 | 17.6 | 5.3 | 10.0 | 24.0 | 15.2 | 3.4 | 9.0 | 22.0 | 14.8 | 3.7 | 9.0 | 21.0 |
| *Impulse regulation* [c] | 23.1 | 7.5 | 13.0 | 40.0 | 23.3 | 8.5 | 14.0 | 39.0 | 17.4 | 4.3 | 12.0 | 29.0 | 15.2 | 3.0 | 11.0 | 21.0 |
| *Social Insecurity* [d] | 23.4 | 6.5 | 14.0 | 35.0 | 20.7 | 6.0 | 14.0 | 32.0 | 18.2 | 3.5 | 10.0 | 27.0 | 17.1 | 3.9 | 12.0 | 21.5 |
| **Body image (Drive for Muscularity Scale & Body Morphing Tool)** | | | | | | | | | | | | | | | | |
| *Drive for muscularity* [e] | 32.8 | 10.4 | 19.0 | 55.0 | 43.1 | 14.2 | 16.0 | 59.0 | 26.8 | 6.7 | 19.0 | 43.0 | 27.6 | 5.6 | 21.0 | 37.0 |
| *Perceived body fat* [a] | -6.7 | 26.5 | -74.8 | 31.7 | -11.7 | 35.5 | -54.7 | 71.9 | -6.6 | 19.9 | -64.8 | 18.0 | -4.5 | 22.1 | -32.7 | 34.5 |
| *Desired body fat* [a] | -31.2 | 26.6 | -79.9 | 2.2 | -18.2 | 52.5 | -81.3 | 100.0 | -22.5 | 24.8 | -92.1 | 3.6 | -23.8 | 24.1 | -71.4 | 7.9 |
| *Perceived muscularity* [a] | 34.5 | 19.4 | .0 | 66.5 | 48.2 | 26.0 | 8.3 | 80.9 | 37.2 | 15.8 | 6.5 | 76.6 | 20.2 | 11.5 | .0 | 35.9 |
| *Desired muscularity* [a] | 42.7 | 19.7 | .0 | 88.5 | 73.4 | 24.0 | 33.2 | 100.0 | 46.9 | 13.8 | 16.1 | 75.1 | 46.2 | 16.6 | 12.4 | 68.2 |
| *Perceived definition* [a] | 38.3 | 14.2 | 6.1 | 60.1 | 70.3 | 20.0 | 28.1 | 100.0 | 39.8 | 14.8 | 7.9 | 71.4 | 39.2 | 19.9 | 7.6 | 65.5 |
| *Desired definition* [a] | 49.3 | 20.2 | 11.9 | 97.5 | 88.4 | 19.7 | 48.9 | 100.0 | 54.8 | 16.5 | 7.4 | 78.4 | 72.9 | 15.3 | 47.9 | 100.0 |
| **Hormonal Status** | | | | | | | | | | | | | | | | |
| *Cortisol [ng/ml]* | 132.9 | 79.7 | 32.7 | 347.9 | 105.6 | 52.3 | 34.2 | 190.1 | 205.5 | 98.0 | 59.9 | 460.8 | 110.0 | 45.6 | 42.6 | 156.4 |
| *Leptin [ng/ml]* | 11.8 | 7.4 | 1.1 | 26.0 | 1.5 | .8 | .5 | 2.8 | 10.6 | 6.4 | 2.5 | 25.6 | 3.2 | 2.5 | .4 | 9.4 |
| *Free T3 [pg/ml]* | 3.5 | 1.5 | 2.1 | 9.6 | 3.2 | .8 | 2.0 | 4.7 | 3.1 | .5 | 2.1 | 4.3 | 3.0 | .6 | 2.0 | 3.7 |
| *LH [mlU/ml]* [h] | 9.4 | 8.6 | .6 | 27.3 | 8.8 | 5.8 | .1 | 18.3 | 13.2 | 11.1 | 1.8 | 34.8 | 9.5 | 5.8 | 3.1 | 19.3 |
| *FSH [mlU/ml]* [h] | 4.8 | 2.9 | .0 | 11.2 | 3.4 | 3.5 | .0 | 12.1 | 5.6 | 3.0 | 1.3 | 10.4 | 3.2 | 2.5 | .7 | 8.0 |
| *Progesterone [ng/ml]* [h] | 2.8 | 4.6 | .3 | 15.8 | .7 | .3 | .4 | 1.2 | 1.9 | 2.8 | .5 | 7.9 | .7 | .3 | .4 | 1.5 |
| *Estradiol [pg/ml]* [h] | 109.0 | 60.5 | 38.4 | 265.5 | 73.6 | 15.0 | 47.4 | 91.3 | 74.1 | 14.3 | 53.8 | 95.6 | 57.9 | 21.5 | 12.3 | 78.3 |
| *Testosterone [ng/ml]* [h] | 1.0 | .3 | .7 | 1.7 | 5.5 | 1.2 | 3.8 | 7.8 | .9 | .2 | .6 | 1.4 | 6.4 | 1.6 | 3.8 | 9.7 |

**Table S2: Groupwise scores/levels of eating disorder psychopathology, hormonal status, and body image assessments**

SD = standard deviation, min = minimum, max = maximum, Missing values: a: n = 1 fHC. b: n = 2 fHC. c: n = 3 fHC. d: n = 4 fHC. e: n = 2 fAN, 3 fHC, 2 mAN. f: n = 2 fAN, 3 fHC. g: n = 1 mHC.

| | Analyses of covariance | | | | | | | | | | | | | Simple main effects | | | |
|---|---|---|---|---|---|---|---|---|---|---|---|---|---|---|---|---|---|
| | Full model | | | | Diagnosis | | | Sex | | | Interaction | | | Female | | Male | |
| | *df* | *F* | *p* | $\eta_p^2$ | *F* | *p* | $\eta_p^2$ | *F* | *p* | $\eta_p^2$ | *F* | *p* | $\eta_p^2$ | *Comp.* | *p* | *Comp.* | *p* |
| **Residual eating disorder psychopathology (Eating Disorder Inventory 2)** | | | | | | | | | | | | | | | | | |
| *Drive for thinness* [a] | 5, 62 | **6.73** | **<.001*** | **.35** | **6.77** | **.012*** | **.10** | **6.07** | **.016*** | **.09** | 3.42 | .069 | .05 | AN>HC | <.001* | ns. | .488 |
| *Bulimia* [a] | 5, 62 | 1.31 | .271 | .10 | .19 | .664 | .00 | 1.46 | .232 | .02 | .78 | .380 | .01 | ns. | .262 | ns. | .688 |
| *Body dissatisfaction* [a] | 5, 62 | **7.98** | **<.001*** | **.39** | **14.77** | **<.001*** | **.19** | **6.78** | **.012*** | **.10** | 1.75 | .190 | .03 | AN>HC | .001* | ns. | .083 |
| *Ineffectiveness* | 5, 63 | **5.85** | **<.001*** | **.32** | **13.95** | **<.001*** | **.18** | 2.07 | .155 | .03 | .83 | .367 | .01 | AN>HC | .001* | ns. | .091 |
| *Perfectionism* [b] | 5, 61 | 2.54 | .038 | .17 | 3.34 | .073 | .05 | .06 | .814 | .00 | 1.17 | .285 | .02 | AN>HC | .022* | ns. | .592 |
| *Interpersonal distrust* [a] | 5, 62 | 2.48 | .041 | .17 | 5.30 | .025* | .08 | .25 | .616 | .01 | .18 | .671 | .00 | AN>HC | .012* | ns. | .275 |
| *Interoceptive awareness* [b] | 5, 61 | **4.46** | **.002*** | **.27** | **10.96** | **.002*** | **.15** | .98 | .327 | .02 | .02 | .899 | .00 | AN>HC | .004* | ns. | .084 |
| *Maturity fears* [a] | 5, 62 | 1.93 | .102 | .14 | .08 | .785 | .00 | 2.24 | .140 | .04 | 1.38 | .245 | .02 | ns. | .213 | ns. | .474 |
| *Asceticism* [c] | 5, 60 | 2.18 | .069 | .15 | 5.45 | .023* | .08 | .31 | .581 | .01 | .47 | .496 | .01 | AN>HC | .012* | ns. | .259 |
| *Impulse regulation* [c] | 5, 60 | **4.72** | **.001*** | **.28** | **15.31** | **<.001*** | **.20** | .01 | .930 | .00 | .33 | .571 | .01 | AN>HC | .003* | AN>HC | .023* |
| *Social Insecurity* [d] | 5, 59 | 3.81 | .005 | .24 | 8.19 | .006* | .12 | .74 | .392 | .01 | .41 | .525 | .01 | AN>HC | .002* | ns. | .166 |
| **Body image (Drive for Muscularity Scale & Body Morphing Tool)** | | | | | | | | | | | | | | | | | |
| *Drive for muscularity* [e] | 5, 56 | **4.18** | **.003*** | **.27** | **15.50** | **<.001*** | **.22** | **4.10** | **.048*** | **.07** | 2.97 | .090 | .05 | AN>HC | .029* | AN>HC | .004* |
| *Perceived body fat* [a] | 5, 62 | 2.96 | .019 | .19 | .10 | .748 | .00 | .04 | .850 | .00 | .41 | .527 | .01 | ns. | .741 | ns. | .703 |
| *Desired body fat* [a] | 5, 62 | .67 | .651 | .05 | .03 | .869 | .00 | .71 | .402 | .01 | .66 | .418 | .01 | ns. | .277 | ns. | .780 |
| *Perceived muscularity* [a] | 5, 62 | 3.42 | .009 | .22 | 8.33 | .005* | .12 | .49 | .489 | .01 | 10.38 | .002* | .14 | ns. | .771 | AN>HC | .001* |
| *Desired muscularity* [a] | 5, 62 | **3.96** | **.004*** | **.24** | **5.08** | **.028*** | **.08** | **9.11** | **.004*** | **.13** | **9.97** | **.002*** | **.14** | ns. | .408 | **AN>HC** | **.002*** |
| *Perceived definition* [a] | 5, 62 | **6.27** | **<.001*** | **.34** | **9.95** | **.002*** | **.14** | **14.84** | **<.001*** | **.19** | **13.20** | **.001*** | **.18** | ns. | .663 | **AN>HC** | **<.001*** |
| *Desired definition* [a] | 5, 62 | **7.66** | **<.001*** | **.38** | .79 | .377 | .01 | **31.09** | **<.001*** | **.33** | **4.47** | **.038*** | **.07** | ns. | .237 | ns. | .071 |
| **Hormonal status** | | | | | | | | | | | | | | | | | |
| *Cortisol* | 5, 63 | **3.89** | **.004*** | **.24** | 2.73 | .103 | .04 | **7.14** | **.010*** | **.10** | 2.38 | .128 | .04 | HC>AN | .004* | ns. | .918 |
| *Leptin* | 5, 63 | **15.31** | **<.001*** | **.55** | <.01 | .948 | .00 | **35.17** | **<.001*** | **.36** | 1.42 | .237 | .02 | ns. | .239 | ns. | .423 |
| *Free T3* | 5, 63 | 1.00 | .423 | .07 | .71 | .404 | .01 | .06 | .805 | .00 | .30 | .584 | .01 | ns. | .301 | ns. | .786 |
| *LH* [f,h] | 5, 33 | 3.02 | .024 | .31 | .02 | .897 | .00 | .20 | .662 | .01 | .23 | .638 | .01 | ns. | .869 | ns. | .428 |
| *FSH* [a,h] | 5, 34 | 2.21 | .076 | .25 | .12 | .736 | .00 | .69 | .413 | .02 | .17 | .685 | .01 | ns. | .654 | ns. | .948 |
| *Progesterone* [h] | 5, 34 | .99 | .440 | .13 | .04 | .844 | .00 | 3.60 | .006 | .01 | .01 | .944 | .00 | ns. | .858 | ns. | .760 |
| *Estradiol* [g,h] | 5, 33 | 2.28 | .069 | .26 | 4.04 | .053 | .11 | 1.22 | .277 | .04 | 1.12 | .301 | .03 | ns. | .099 | ns. | .266 |
| *Testosterone* [h] | 5, 34 | **64.4** | **<.001*** | **.90** | 1.42 | .242 | .04 | **253.67** | **<.001*** | **.88** | **5.63** | **.023*** | **.14** | ns. | .195 | ns. | .063 |

#### Table S3: Group comparison of eating disorder psychopathology, body image assessments and hormonal status

Statistic: two-way analyses of covariance with bootstrapping with inclusion of *diagnosis* and *sex* main effects, the interaction of *diagnosis* × *sex*, and age as well as BMI-SDS as covariates. Significant interaction effects were followed up on by evaluation of simple main effects of diagnosis within each sex category (Šidák correction). Partial eta-squared is used to demonstrate effect sizes. * significant at alpha-level of $p < 0.05$, with Bonferroni correction per set of analyses at the level of full model ($p < .0045$, $p < .0063$, $p < .0071$). Bold numbers indicate effects significant on factor- *and* model-level, and simple-main effects *if* corresponding interaction *and* model effects were significant. Missing values: a: n = 1 fHC. b: n = 2 fHC. c: n = 3 fHC. d: n = 4 fHC. e: n = 2 fAN, 3 fHC, 2 mAN. f: n = 1 fAN, g: n = 1 mHC, h: excluding probands with hormonal contraception. fT3 = free triiodothyronine, LH = luteinizing hormone, FSH = follicle-stimulating hormone, df = degrees of freedom, $\eta_p^2$ = partial eta-squared, comp. = comparison, ns. = not significant.

| | Analyses of covariance | | | | | | | | | | | | | | Simple main effects | | |
|---|---|---|---|---|---|---|---|---|---|---|---|---|---|---|---|---|---|
| | Full model | | | | Diagnosis | | | Sex | | | Interaction | | | Females | | Males | |
| | *df* | *F* | *p* | $\eta_p^2$ | *F* | *p* | $\eta_p^2$ | *F* | *p* | $\eta_p^2$ | *F* | *p* | $\eta_p^2$ | *Comp.* | *p* | *Comp.* | *p* |
| **Residual eating disorder psychopathology (Eating Disorder Inventory 2)** | | | | | | | | | | | | | | | | | |
| *Drive for thinness* [a] | 5, 53 | **6.37** | **<.001*** | **.38** | 3.84 | .055 | .07 | **8.07** | **.006*** | **.13** | 3.57 | .064 | .06 | AN>HC | <.006* | ns. | .963 |
| *Body dissatisfaction* [a] | 5, 53 | **6.73** | **<.001*** | **.39** | **8.57** | **.005*** | **.14** | **7.87** | **.007*** | **.13** | 1.93 | .170 | .04 | AN>HC | .001* | ns. | .369 |
| *Ineffectiveness* | 5, 54 | **4.57** | **.002*** | **.30** | **8.55** | **.005*** | **.14** | 2.65 | .110 | .05 | 1.43 | .237 | .03 | AN>HC | .001* | ns. | .406 |
| *Interoceptive awareness* [a] | 5, 53 | **3.15** | **.015*** | **.23** | **6.07** | **.017*** | **.10** | .93 | .340 | .02 | .16 | .687 | .00 | AN>HC | .012* | ns. | .317 |
| *Impulse regulation* [b] | 5, 52 | **3.17** | **.014*** | **.23** | **9.58** | **.003*** | **.16** | .02 | .656 | .00 | .08 | .776 | .00 | AN>HC | .015* | ns. | .143 |
| **Body image (Drive for Muscularity Scale & Body Morphing Tool)** | | | | | | | | | | | | | | | | | |
| *Drive for muscularity* [c] | 5, 48 | **2.63** | **.035*** | **.22** | **6.66** | **.013*** | **.12** | .83 | .372 | .02 | 1.04 | .312 | .02 | ns. | .131 | AN>HC | .048* |
| *Desired muscularity* [a] | 5, 53 | 1.96 | .100 | .16 | 2.49 | .120 | .05 | 4.75 | .034* | .08 | 5.51 | .023* | .09 | ns. | .431 | ns. | .093 |
| *Perceived definition* [a] | 5, 53 | **3.81** | **.005*** | **.27** | **7.04** | **.011*** | **.12** | **10.28** | **.002*** | **.16** | **7.28** | **.009*** | **.12** | ns. | .955 | **AN>HC** | **.013*** |
| *Desired definition* [a] | 5, 53 | **6.04** | **<.001*** | **.36** | .96 | .331 | .02 | **25.85** | **<.001*** | **.33** | 2.07 | .156 | .04 | ns. | .642 | ns. | .203 |
| **Hormonal status** | | | | | | | | | | | | | | | | | |
| *Cortisol* | 5, 54 | **2.82** | **.025*** | **.21** | 1.35 | .251 | .02 | **5.21** | **.026*** | **.09** | 2.43 | .125 | .04 | HC>AN | .018* | ns. | .701 |
| *Leptin* | 5, 54 | **14.53** | **<.001*** | **.58** | .27 | .605 | .01 | **38.47** | **<.001*** | **.41** | 2.51 | .119 | .04 | ns. | .328 | ns. | .116 |
| *Testosterone* [d] | 5, 29 | **59.92** | **<.001*** | **.91** | .84 | .367 | .03 | **236.50** | **<.001*** | **.89** | 3.26 | .082 | .10 | ns. | .347 | ns. | .130 |

**Table S4: Group comparison results of eating disorder psychopathology, hormonal status, and body image assessments while excluding patients with a BMI below the 10th percentile**

Only main analyses significant at Bonferroni corrected model level (see Table S3) were repeated. No further multiple comparison correction was applied. Group sizes: N = 20 fAN, 7 mAN, 24 fHC, 9 mHC. Statistics: two-way analyses of covariance with bootstrapping with inclusion of diagnosis and sex main effects, the interaction of diagnosis × sex, and age as well as BMI-SDS as covariates. Significant interaction effects were followed up on by evaluation of simple main effects of diagnosis within each sex category (Šidák correction). Partial eta-squared is used to demonstrate effect sizes. * significant at alpha-level of $p < 0.05$, uncorrected. Bold numbers indicate effects significant on factor- and model-level, and simple-main effects if corresponding interaction and model effects were significant. fT3 = free triiodothyronine, LH = luteinizing hormone, FSH = follicle-stimulating hormone, df = degrees of freedom, ηp2 = partial eta-squared, comp. = comparison, ns. = not significant, missing values: a: n = 1 fHC, b: n =2 fHC, c: n =1 fAN, n = 3 fHC, n = 2 mHC.

| | Analyses of covariance | | | | | | | | | | | | | Simple main effects | | | |
|---|---|---|---|---|---|---|---|---|---|---|---|---|---|---|---|---|---|
| | **Full model** | | | | **Diagnosis** | | | **Sex** | | | **Interaction** | | | **Females** | | **Males** | |
| | *df* | *F* | *p* | $\eta_p^2$ | *F* | *p* | $\eta_p^2$ | *F* | *p* | $\eta_p^2$ | *F* | *p* | $\eta_p^2$ | *Comp.* | *p* | *Comp.* | *p* |
| **Residual eating disorder psychopathology (Eating Disorder Inventory 2)** | | | | | | | | | | | | | | | | | |
| *Drive for thinness* [a] | 5, 65 | **6.14** | **<.001*** | **.32** | **10.34** | **.002*** | **.14** | 3.53 | .065 | .05 | 1.63 | .206 | .03 | AN>HC | <.001* | ns. | .254 |
| *Body dissatisfaction* [a] | 5, 65 | **8.35** | **<.001*** | **.39** | **19.13** | **<.001*** | **.23** | **4.88** | **.031*** | **.07** | 1.01 | .319 | .02 | AN>HC | <.001* | ns. | .050 |
| *Ineffectiveness* | 5, 66 | **6.50** | **<.001*** | **.33** | **17.98** | **<.001*** | **.21** | 1.65 | .203 | .02 | .47 | .496 | .01 | AN>HC | <.001* | AN>HC | .040* |
| *Interoceptive awareness* [b] | 5, 64 | **5.19** | **<.001*** | **.29** | **14.72** | **<.001*** | **.19** | .60 | .440 | .01 | .00 | .996 | .00 | AN>HC | .001* | AN>HC | .025* |
| *Impulse regulation* [c] | 5, 63 | **5.59** | **<.001*** | **.31** | **18.16** | **<.001*** | **.22** | .00 | .955 | .00 | .17 | .684 | .00 | AN>HC | .001* | AN>HC | .007* |
| **Body image (Drive for Muscularity Scale & Body Morphing Tool)** | | | | | | | | | | | | | | | | | |
| *Drive for muscularity* [d] | 5, 59 | **4.60** | **.001*** | **.28** | **17.56** | **<.001*** | **.23** | 3.54 | .065 | .06 | 2.90 | .094 | .05 | AN>HC | .020* | AN>HC | .001* |
| *Desired muscularity* [a] | 5, 65 | **5.17** | **<.001*** | **.29** | **5.62** | **.021*** | **.08** | **10.46** | **.002*** | **.14** | **12.54** | **.001*** | **.16** | ns. | .285 | **AN>HC** | **.001*** |
| *Perceived definition* [a] | 5, 65 | **4.72** | **.001*** | **.27** | **5.72** | **.020*** | **.08** | **11.35** | **.001*** | **.15** | **10.03** | **.002*** | **.13** | ns. | .483 | **AN>HC** | **.002*** |
| *Desired definition* [a] | 5, 65 | **9.07** | **.001*** | **.41** | 0.76 | .388 | .01 | **33.86** | **<.001*** | **.34** | **4.99** | **.029*** | **.07** | ns. | .207 | ns. | .073 |
| **Hormonal status** | | | | | | | | | | | | | | | | | |
| *Cortisol* | 5, 66 | **3.49** | **.007*** | **.21** | 2.09 | .153 | .03 | **6.41** | **.014*** | **.09** | 1.86 | .177 | .03 | HC>AN | .009* | ns. | .957 |
| *Leptin* | 5, 66 | **13.80** | **<.001*** | **.51** | .04 | .834 | .00 | **29.96** | **<.001*** | **.31** | .48 | .490 | .01 | ns. | .393 | ns. | .779 |
| *Testosterone* [e] | 5, 36 | **69.55** | **<.001*** | **.91** | 1.40 | .244 | .04 | **267.27** | **<.001*** | **.88** | **7.13** | **.011*** | **.17** | ns. | .120 | **HC>AN** | **.047*** |

**Table S5: Group comparison results of eating disorder psychopathology, hormonal status, and body image assessments while additionally including subjects with current Bulimia nervosa**

Only main analyses significant at Bonferroni corrected model level (see Table S3) were repeated. No further multiple comparison correction was applied. Group sizes: N = 26 fAN, 10 mAN, 26 fHC, 10 mHC. Statistics: two-way analyses of covariance with bootstrapping with inclusion of diagnosis and sex main effects, the interaction of diagnosis × sex, and age as well as BMI-SDS as covariates. Significant interaction effects were followed up on by evaluation of simple main effects of diagnosis within each sex category (Šidák correction). Partial eta-squared is used to demonstrate effect sizes. * significant at alpha-level of $p < 0.05$, uncorrected. Bold numbers indicate effects significant on factor- and model-level, and simple-main effects if corresponding interaction and model effects were significant. fT3 = free triiodothyronine, LH = luteinizing hormone, FSH = follicle-stimulating hormone, df = degrees of freedom, ηp2 = partial eta-squared, comp. = comparison, ns. = not significant, missing values: a: n = 1 fHC, b: n = 2 fHC, c: n = 3 fHC, d: n = 2 fAN, n = 3 fHC, n = 2 mHC.

| | Mann-Whitney-U tests | | | | | | | | | | | |
|---|---|---|---|---|---|---|---|---|---|---|---|---|
| | HC vs. AN | | | Male vs. Female | | | fAN vs. fHC | | | mAN vs. mHC | | |
| | *N* | *U* | *p* | *N* | *U* | *p* | *N* | *U* | *p* | *N* | *U* | *p* |
| **Residual eating disorder psychopathology (Eating Disorder Inventory 2)** | | | | | | | | | | | | |
| *Drive for thinness* | 68 | **840.5** | **.001*** | 68 | **305.5** | **.028*** | 49 | **461.5** | **.001*** | 19 | 55.0 | .410 |
| *Body dissatisfaction* | 68 | **921.5** | **<.001*** | 68 | **290.0** | **.016*** | 49 | **495.0** | **<.001*** | 19 | 68.5 | .054 |
| *Ineffectiveness* | 69 | **929.5** | **<.001*** | 69 | **321.5** | **.039*** | 50 | **498.5** | **<.001*** | 19 | 67.0 | .072 |
| *Interoceptive awareness* | 67 | **852.0** | **<.001*** | 67 | 319.0 | .056 | 48 | **441.5** | **.002*** | 19 | 66.0 | .085 |
| *Impulse regulation* | 66 | **860.5** | **<.001*** | 66 | 365.0 | .250 | 47 | **423.0** | **.002*** | 19 | **75.5** | **.013*** |
| **Body image (Drive for Muscularity Scale & interactive Body Morphing Tool)** | | | | | | | | | | | | |
| *Drive for muscularity* | 62 | **682.0** | **.005*** | 62 | 487.0 | .099 | 45 | 336.5 | .058 | 17 | **61.0** | **.016*** |
| *Desired muscularity* | 68 | 623.0 | .577 | 68 | **625.5** | **.029*** | 49 | 265.5 | .490 | 19 | **71.0** | **.034*** |
| *Perceived definition* | 68 | 547.5 | .713 | 68 | **650.0** | **.012*** | 49 | 285.0 | .764 | 19 | **79.0** | **0.06*** |
| *Desired definition* | 68 | 678.5 | .215 | 68 | **785.5** | **<.001*** | 49 | 239.5 | .226 | 19 | 68.5 | .051 |
| **Hormonal status** | | | | | | | | | | | | |
| *Cortisol* | 69 | **367.5** | **.007*** | **69** | **285.5** | **.011*** | 50 | **159.5** | **.003*** | 19 | 43.0 | .870 |
| *Leptin* | 69 | 579.0 | .857 | **69** | **60.0** | **<.001*** | 50 | 338.5 | .607 | 19 | 23.5 | .079 |
| *Testosterone* | 40 | 156.5 | .287 | **40** | **399.0** | **<.001*** | 21 | 33.0 | .255 | 19 | 28.5 | .178 |

**Table S6: Replication of significant group comparisons using Mann-Whitney U tests**

Only main analyses significant at Bonferroni corrected model level (see Table S3) were repeated. * significant at an alpha-level of p < .05, uncorrected. U = Mann-Whitney U statistic.

| Variable 1 | Variable 2 | Recovered AN | | Healthy controls | |
|---|---|---|---|---|---|
| | | Female | Male | Female | Male |
| | | *rs, p (N)* | *rs, p (N)* | *rs, p (N)* | *rs, p (N)* |
| ***Perceived body fat*** | *BMI-SDS* | **.71, <.001* (24)** | .83, .831 (9) | **.61, .001* (25)** | .20, .580 (10) |
| | *Leptin* | **.43, .039* (24)** | .65, .058 (9) | **.58, .002* (25)** | .28, .434 (10) |
| | *DMS* | -.39, .075 (22) | -.37, .330 (9) | .27, .221 (22) | -.17, .686 (8) |
| | *Drive for thinness* | **.47, .021* (24)** | .16, .680 (9) | **.74, <.001* (24)** | .46, .186 (10) |
| ***Desired body fat*** | *BMI-SDS* | **.47, .020* (24)** | -.22, .576 (9) | -.16, .454 (25) | .29, .425 (10) |
| | *Leptin* | **.47, .021* (24)** | -.43, .244 (9) | -.15, .483 (25) | -.28, .434 (10) |
| | *DMS* | **-.45, .036* (22)** | -.43, .252 (9) | **-.71, <.001 (22)** | -.68, .062 (8) |
| | *Drive for thinness* | -.28, .192 (24) | .34, .374 (9) | -.28, .184 (24) | .62, .058 (10) |
| ***Perceived muscularity*** | *BMI-SDS* | -.22, .313 (24) | **.90, .001* (9)** | .35, .082 (25) | -.04, .907 (10) |
| | *Leptin* | -.26, .229 (24) | -.02, .996 (9) | .05, .815 (25) | -.15, .688 (10) |
| | *DMS* | -.12, .592 (22) | -.08, .831 (9) | .30, .169 (22) | -.17, .686 (8) |
| | *Drive for thinness* | -.20, .342 (24) | **-.77, .016* (9)** | .31, .146 (24) | -.27, .449 (10) |
| ***Desired muscularity*** | *BMI-SDS* | -.30, .154 (24) | .23, .559 (9) | .21, .325 (25) | .03, .934 (10) |
| | *Leptin* | -.18, .396 (24) | -.39, .306 (9) | .21, .327 (25) | .26, .466 (10) |
| | *DMS* | .16, .482 (22) | **.68*, .045* (9)** | .02, .932 (22) | .20, .643 (8) |
| | *Drive for thinness* | .03, .878 (24) | .17, .655 (9) | .10, .646 (24) | -.10, .774 (10) |
| ***Perceived definition*** | *BMI-SDS* | -.19, .371 (24) | .05, .898 (9) | .22, .293 (25) | .53, .117 (10) |
| | *Leptin* | **-.50, .013* (24)** | -.33, .381 (9) | -.27, .196 (25) | -.54, .111 (10) |
| | *DMS* | .08, .729 (22) | .02, .966 (9) | -.31, .159 (22) | .24, .560 (8) |
| | *Drive for thinness* | -.13, .551 (24) | .00, 1.00 (9) | .10, .627 (24) | .50, .137 (10) |
| ***Desired definition*** | *BMI-SDS* | -.23, .388 (24) | -.21, .604 (9) | -.11, .589 (25) | .48, .166 (10) |
| | *Leptin* | -.06, .791 (24) | -.46, .217 (9) | -.09, .655 (25) | -.38, .275 (10) |
| | *DMS* | .21, .350 (22) | .24, .537 (9) | -.54, .811 (22) | .00, 1.000 (8) |
| | *Drive for thinness* | -.06, .791 (24) | **.83, .005* (9)** | -.15, .493 (24) | .38, .285 (10) |

**Table S7: Correlational analyses to validate the Body Morphing Tool results**

Variables were correlated using spearman correlations. * significant at uncorrected alpha-level of p < .05. AN = Anorexia nervosa, rs = Spearman correlation coefficient, BMI-SDS = body mass index – standard deviation score, DMS = Drive for Muscularity Scale.

#### Supplementary Figures

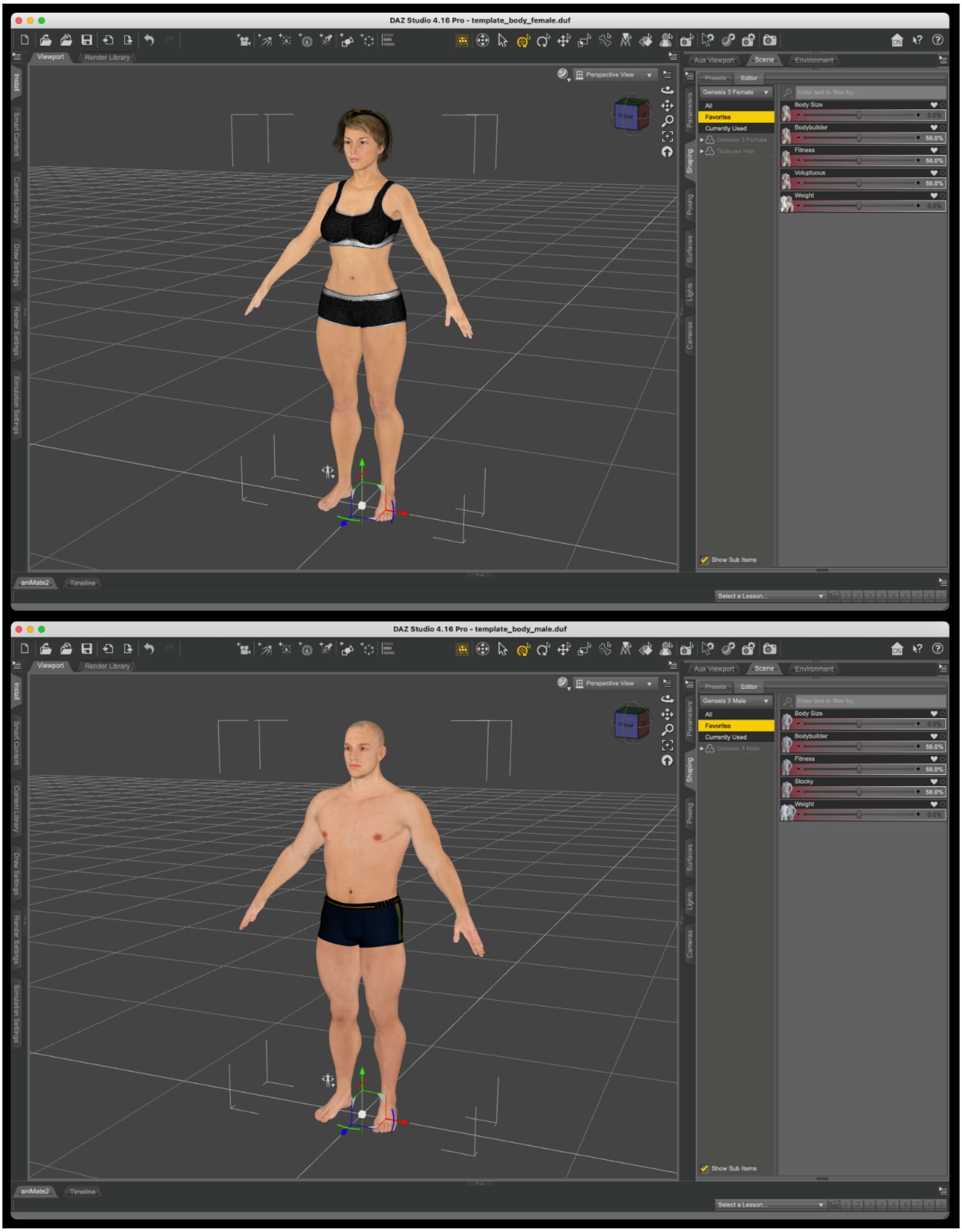

**Figure S1: User interface of the Body Morphing Tool (BMT)**

Software: *DAZ Studio 4 Pro* (https://www.daz3d.com/). The templates were built from packages *Genesis 3 Starter Essentials*, *Female* and *Male Body Morphs*. Participants could move the camera freely around the bodies. For simplicity, five parameters were selected before the experiment to transform the bodies into the desired shape (right side).

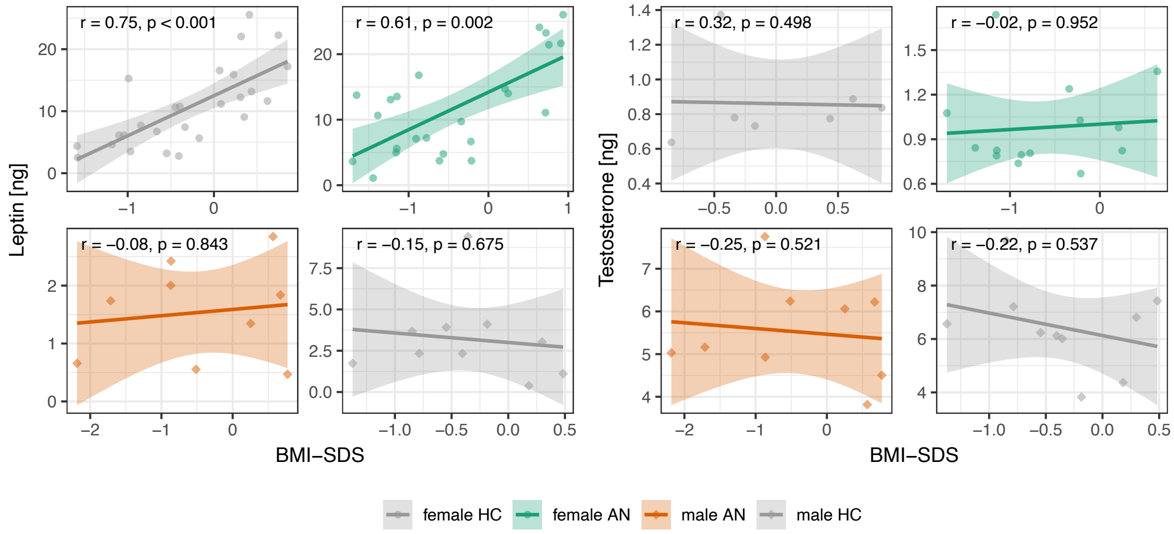

**Figure S2: Correlation of leptin and testosterone with BMI-SDS**

Spearman correlations were conducted separately by group. Scatter circles and squares depict individual values, regression lines and 95% confidence intervals are shown. Leptin is measured in ng/ml. AN = Anorexia nervosa, HC = healthy controls, BMI-SDS = body mass index – standard deviation score.

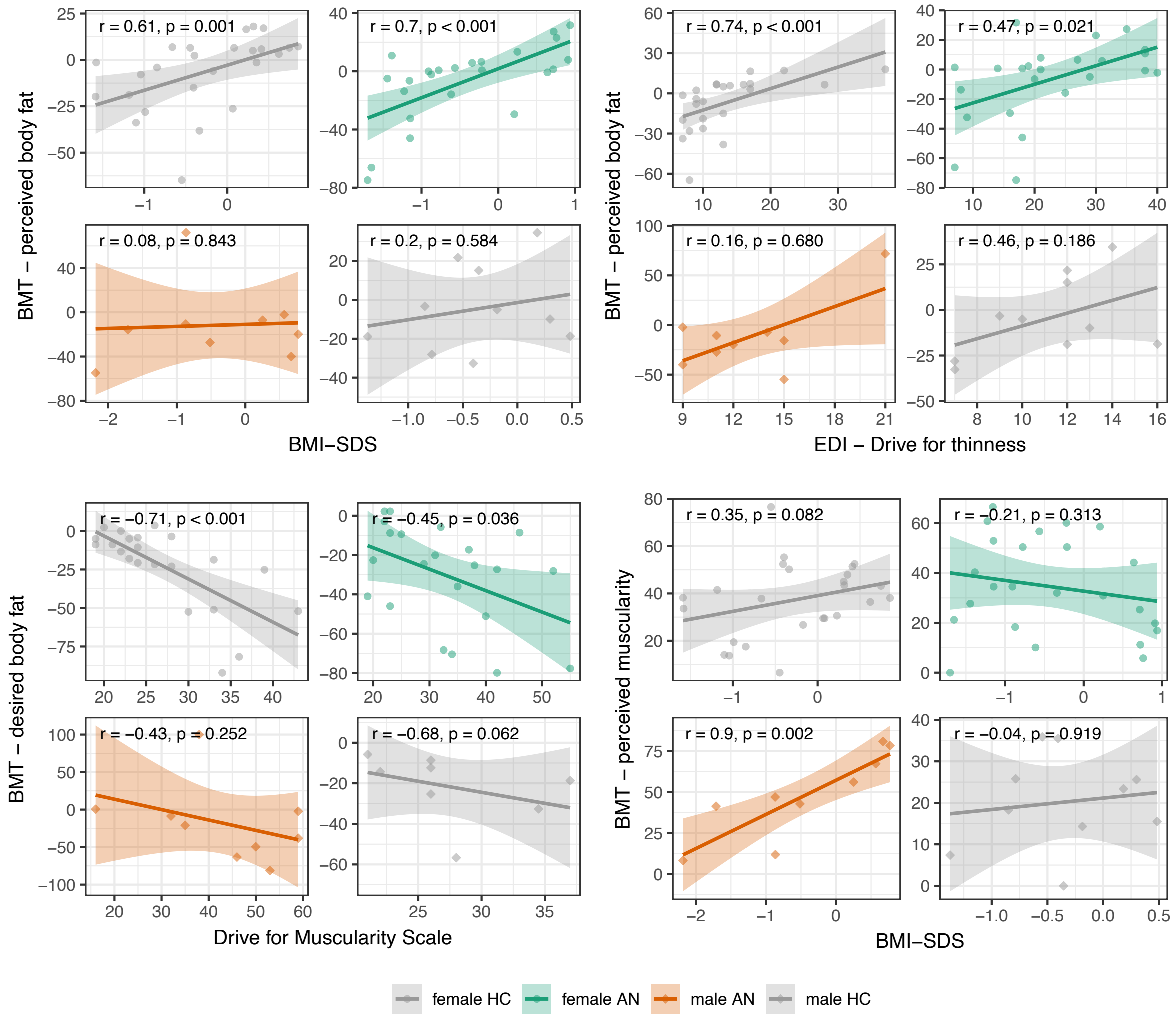

**Figure S3: Exemplary associations between body weight, drive for muscularity, drive for thinness, and Body Morphing Tool results**

Spearman correlations were conducted separately by group. Scatter circles and squares depict individual values, regression lines and 95% confidence intervals are shown. See Table S5 for full results. AN = Anorexia nervosa, HC = Healthy controls, BMT = Body Morphing Tool, BMI-SDS = body mass index – standard deviation score, EDI = Eating Disorder Inventory 2.
